# Salivary anti-SARS-CoV-2 IgA as an accessible biomarker of mucosal immunity against COVID-19

**DOI:** 10.1101/2020.08.07.20170258

**Authors:** Atul Varadhachary, Dev Chatterjee, Javier Garza, R. Patrick Garr, Christopher Foley, Andrea Letkeman, John Dean, David Haug, Juliet Breeze, Robbyn Traylor, Andrew Malek, Rohan Nath, Leo Linbeck

**Affiliations:** BreviTest Technologies, LLC and Fannin Innovation Studio, Houston, TX; Arctas Capital Group, Houston, TX; Next Level Urgent Care, Houston, TX; Alchemy Industrial, LLC, Houston, TX; APC Health, Pearland, TX

## Abstract

**Background:** Mucosal immunity, including secretory IgA (sIgA), plays an important role in early defenses against respiratory pathogens. Salivary testing, the most convenient way to measure sIgA, has been used to characterize mucosal immune responses to many viral infections including SARS, MERS, influenza, HIV, and RSV. However, its role has not yet been characterized in the COVID-19 pandemic. Here, we report development and validation of a rapid immunoassay for measuring salivary IgA against the SARS-CoV-2 virus, and report quantitative results in both pre-COVID-19 and muco-converted subjects.

**Methods:** We developed and refined a specific test for salivary IgA against SARS-CoV-2 on the Brevitest platform, a rapid immunoassay system designed for point-of-care use. A qualitative test was validated as per FDA guidelines with saliva obtained from subjects prior to the emergence of COVID-19, and from PCR-confirmed COVID-19 patients. We also generated a quantitative measure of anti-SARS-CoV-2 salivary IgA. Time taken for saliva self-collection was measured and its ease-of-use assessed.

**Results:** We successfully validated a qualitative salivary assay for SARS-CoV-2 IgA antibodies, with positive and negative predictive values of 92% and 97%, respectively, and no observable cross-reactivity with any of seven potential confounders. Pre-COVID-19 saliva samples showed an 8-fold range of IgA concentrations, suggesting a broad continuum of natural antibody resistance against the novel virus, though at levels lower than that observed in COVID-19 PCR-confirmed subjects. Samples from muco-positive subjects also shown a ~9-fold variation in salivary IgA levels, with elevated salivary IgA observed beyond three months after onset of symptoms. We observed a correlation (r=0.4405) between salivary IgA levels and COVID-19 disease severity. In anecdotal observations, we observed individuals who exhibited antibodies early in the course of their disease, contemporaneously with a positive PCR test, as well as individuals who muco-converted despite no known direct exposure to a COVID-19 patient, no symptoms, and negative molecular and/or serum antibody tests. Salivary collection took 5-10 minutes, and was reported as being easy (mean of 1.1 on a scale of 1 to 10).

**Implications:** Mucosal immunity, including secretory IgA, plays an important role in host defense against respiratory pathogens, and our early data suggest it may do so in COVID-19. Salivary IgA, an accessible marker of mucosal immunity, may be a useful indicator of several key parameters including individual and community immune response, disease severity, clinical risk, and herd immunity. The non-invasive nature and ease of saliva collection facilitates its potential use as a biomarker for ongoing patient assessment and management, as well as a community surveillance tool. By measuring mucosal immune responses directly and systemic immune responses indirectly, salivary IgA could be useful in developing and deploying a vaccine(s) against COVID-19. Quantitative IgA assessment could also potentially serve as a tool to segment the population into different risk categories and inform individual and collective decisions relating to appropriate activities and vaccine prioritization/delivery. These data reinforce the importance of further investigation into the role of mucosal immunity and IgA in host responses against COVID-19.

## INTRODUCTION

The COVID-19 pandemic has been characterized by rapid global spread and has impacted the life of almost every person on the planet. First reported in the Wuhan province in China in December 2019, the COVID-19 disease reached pandemic status within six months and has spread to nearly every country. Although initially contained in several countries, COVID-19 has begun to resurface even as it continues to surge through other countries, such as the United States, Russia, India and Brazil, which have had less success with containment or are experiencing rapid increases in the number of cases^1,2^. COVID-19 is caused by a novel coronavirus, termed severe acute respiratory syndrome coronavirus 2 (SARS-CoV-2) by the World Health Organization in February 2020^3^. Coronaviruses have been responsible for several respiratory disease outbreaks over the previous two decades, including Severe Acute Respiratory Syndrome (SARS, caused by the SARS-CoV-1 virus) which was primarily limited to Southeast Asia, and Middle East Respiratory Syndrome (MERS, caused by the MERS-CoV). Although the exact mechanisms behind the increased spread of SARS-CoV-2 remains to be discovered, one hypothesis suggests that SARS-CoV-2 spread is fueled by the infectivity of asymptomatic or pre-symptomatic carriers, making containment difficult and allowing the virus to spread worldwide through travel and community-based contacts^4,5^.

SARS-CoV-2 appears to be primarily spread via respiratory droplets which begin as mucosal secretions in infected individuals. These droplets become aerosolized by coughing, sneezing, or talking and can spread through the air or through contaminating surfaces. Respiratory droplets are particularly infectious when infected persons are in enclosed areas or in close contact with others^6^. Compounding the challenges of preventing transmission of the disease, symptoms can vary widely in severity; some patients remain largely asymptomatic or present with mild disease while others may develop a potentially fatal severe respiratory crisis^7^. Common symptoms include sore throat, fever, cough, muscle pain, headache, and a characteristic loss of taste or smell. Severe cases may result in progressive lung pathology beginning with difficulty breathing, and progressing to pneumonia or acute respiratory distress syndrome (ARDS), often requiring intubation and mechanical ventilation of the lungs^8^. ARDS is typically associated with a cytokine storm, and can result in organ damage and end stage failure in organs beyond the lung, including the brain, gastrointestinal tract, vasculature, kidney, liver and heart^9^,^10^.

Outside of efforts to limit physical exposure, including wearing face coverings and limiting contact with others, the first line of defense against a viral respiratory infection is the mucosal immune system in the respiratory tract. The mucosal immune system plays critical roles in both innate and adaptive immunity. As part of the innate response, the mucosal immune system coats the lumen of the respiratory tract with a liquid lining containing surfactants, mucus, and periciliary fluid. A rapid response to a pathogen challenge to the respiratory tract is triggered by ‘sensor’ cells, which include epithelial cells, macrophages, dendritic cells, and mast cells. Upon detection of a pathogen, these cells trigger innate responses, which include the generation of reactive oxygen species and anti-microbial peptides^11,12,13,14^ targeted against the pathogen, along with the release of cytokines which attract neutrophils, natural killer cells, and lymphoid cells. In conjunction with mucociliary clearance, and, where appropriate, recruitment of additional responder cells (such as eosinophils, basophils, and monocytes), the mucosal innate immune system protects the respiratory tract and facilitates pathogen clearance.

When the viral inoculum is large enough, it can trigger participation by the mucosal adaptive immune system, mediated in part through dendritic cells – protruding through the epithelium – to sample luminal contents^15, 16, 17^. Dendritic cells, play a critical role in activating an adaptive response.

Part of this adaptive response is the generation of antibodies against the pathogen, including IgA which plays an important role in mucosal immunity. IgA, which is the antibody class produced in largest quantities by the body^18^ primarily exists in the body in two forms: the predominant polymeric form is a dimer (two IgA molecules covalently linked along the Fc region via J chain) found in mucosal secretions, while the less abundantly monomeric form is primarily found in serum. Plasma cells producing dimeric IgA localize along the lamina propria adjacent to mucosal membranes. Dimeric IgA is translocated across the epithelium via the polymeric immunoglobulin receptor^18^ and is secreted into the mucosa, where the covalent linkage confers protease resistance to the IgA molecule^17,19^. This secretory IgA (sIgA) has a number of essential functions within mucosal immunity. A major function is to prevent the cognate pathogens from infecting hos cells via immune exclusion, by competing for the host-cell ligands that trigger viral entry. sIgA further contributes to viral clearance via agglutination and shielding of microbial adhesins for later clearance via ciliary activity^20^. In the case of SARS-CoV-2, sIgA antibodies may prevent adhesion to target epithelial cells via neutralization of the coronavirus spike protein (and thus inhibiting its interaction with the host ACE-2 receptor^21^) or binding to the SARS-CoV-2 nucleocapsid protein. Beyond its role in immune exclusion, sIgA can initiate and regulate the process of myeloid immune responses through the FcaR receptor to the IgA FC region that is found on multiple immune and epithelial cells, resulting in a broad range of effector functions involving both humoral and cellular responses. The production of IgA, relative to other antibody classes is closely controlled by the class-switching process.

IgA responses have already been described as playing a role in immune responses to SARS-CoV-2. Serum IgA has been detected in COVID-19 patients and appears to be detectable earlier than IgM or IgG antibodies^22^, possibly as early as two days after onset of symptoms (versus five days for either IgM or IgG)^23^. This suggests that IgA may be the first antibody to appear in response to SARS-CoV-2 infection, but research into the role of IgA, especially mucosal IgA in COVID-19 has lagged.

Beyond its important role in mucosal immunity and its clear importance in regulating host response to respiratory infections, IgA is attractive for public health response for another reason: its localization to the mucosa. As a secreted antibody, sIgA is easily accessible in saliva, and compared with other sample types, multiple groups have recognized the attractiveness of salivary testing due to its ease of collection^24,25.^

Here we report results from over 250 tests IgA tests, including those from earlier versions of our assay. The IgA test was developed using the Brevitest platform, which is designed for rapid, quantitative, point-of-care ELISA testing. The test was validated for qualitative detection of IgA in saliva in accordance with FDA guidelines using PCR-confirmed subjects as the true positive group and saliva samples obtained before COVID-19 emergence as true negative. In addition, we share quantitative data on levels of salivary IgA against SARS-CoV-2 in both pre-COVID and muco-positive samples. These analyses demonstrate a broad range of IgA concentrations across subjects, suggest the persistence of anti-SARS-CoV-2 salivary IgA through at least 3 months after onset of symptoms and a potential correlation between salivary IgA levels and disease severity. We also present data on time and ease of use in self-collected saliva samples. Our findings suggest a potential role for salivary IgA testing in managing individual patients, understanding community infection rates, and supporting vaccine development and deployment.

## RESULTS

We utilized our Brevitest platform to develop and validate our salivary IgA assay, which we term BRAVO (Brevitest IgA Salivary Mucosal Test). Briefly, the Brevitest platform is a rapid, automated enzyme-linked immunosorbent assay (ELISA) designed for point-of-care (POC) use. As the platform is automated, minimal variation between loading, reagents, washing, and development times are expected. Additional details regarding the platform can be found in the Methods. We developed our COVID-19 salivary IgA assay to detect IgA antibodies against either the SARS-CoV-2 Spike S1 antigen (S1) or the SARS-CoV-2 Nucleocapsid Protein (NP) antigen. To validate the BRAVO assay and to ensure that we were detecting antibodies within a linear range, we spiked increasing concentrations of commercially available anti-S1 and anti-NP antibodies into simulated saliva. We observed a strong, linear correlation in our BRAVO assay (**Figure 1**) with a p-value of <0.0001 calculated with a simple linear regression.

**Figure 1.**
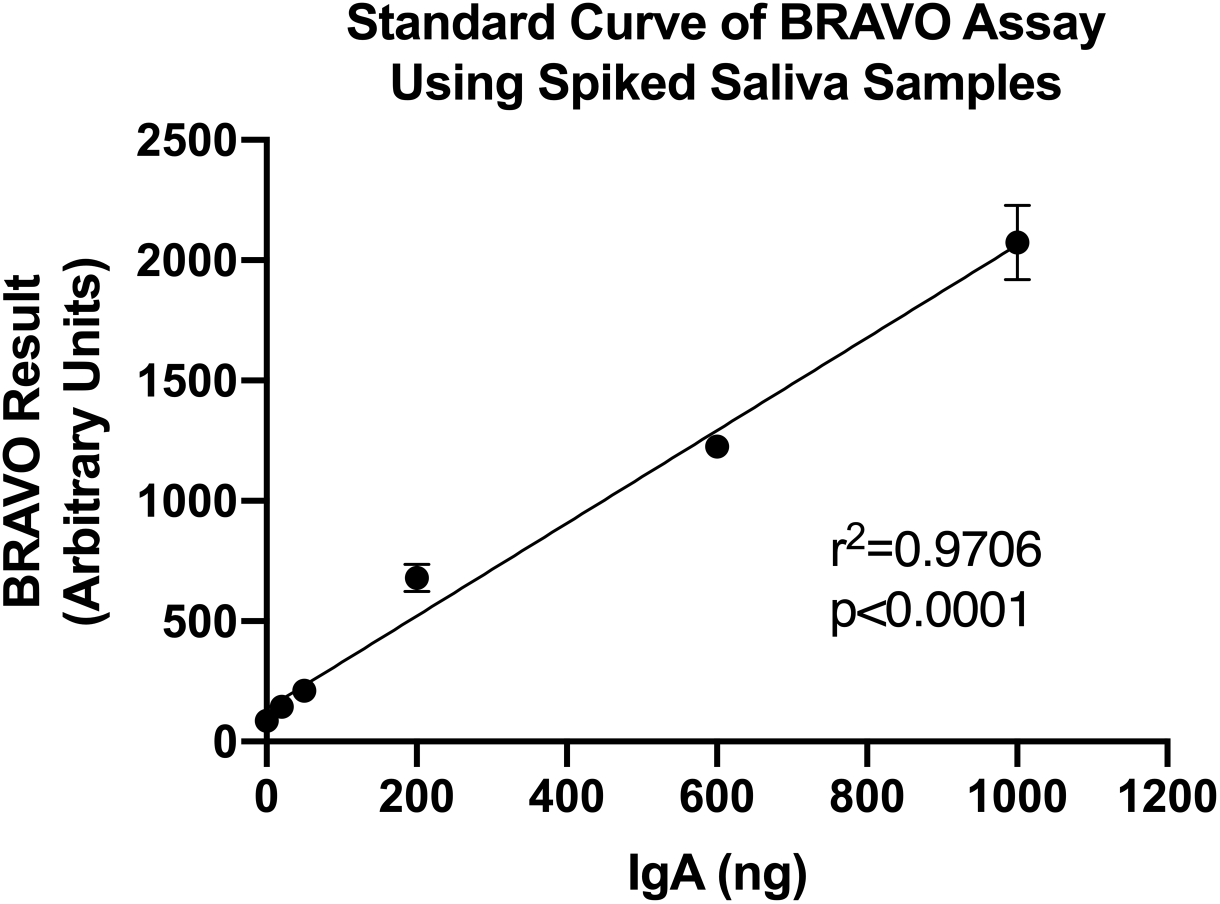
Increasing concentrations of commercially available anti-S1 and anti-NP IgA antibodies were spiked into simulated saliva in order to mimic COVID-19 patient samples. These samples were loaded onto the BRAVO assay and run via the Brevitest analyzer. Results indicated a broad linear range of acceptable antibody concentrations that could be robustly detected with BRAVO assay. A simple linear regression was utilized to demonstrate the linear range of the BRAVO assay (r^2^=0.9706, p<0.0001).

After validating the linear range and parameters of the BRAVO assay, we next sought to determine a cut-off threshold for positive and negative sample types. We purchased true negative samples (saliva collected pre-COVID-19) to determine our negative mean. Interestingly, we observed significant variation (~9-fold) in salivary IgA levels among the true negative samples (**Figure 2**), potentially explained by the presence of innate, polyreactive IgA or antibodies against other strains of coronavirus. The mean of our negative sample cohort was determined to be 43.2 Arbitrary Units (AU), with a standard deviation of 19.4 AU. We selected a value at which 95% of the samples returned the expected negative result, which resulted in a threshold cut-off of 1.65 standard deviations above the mean, or 75 AU.

**Figure 2.**
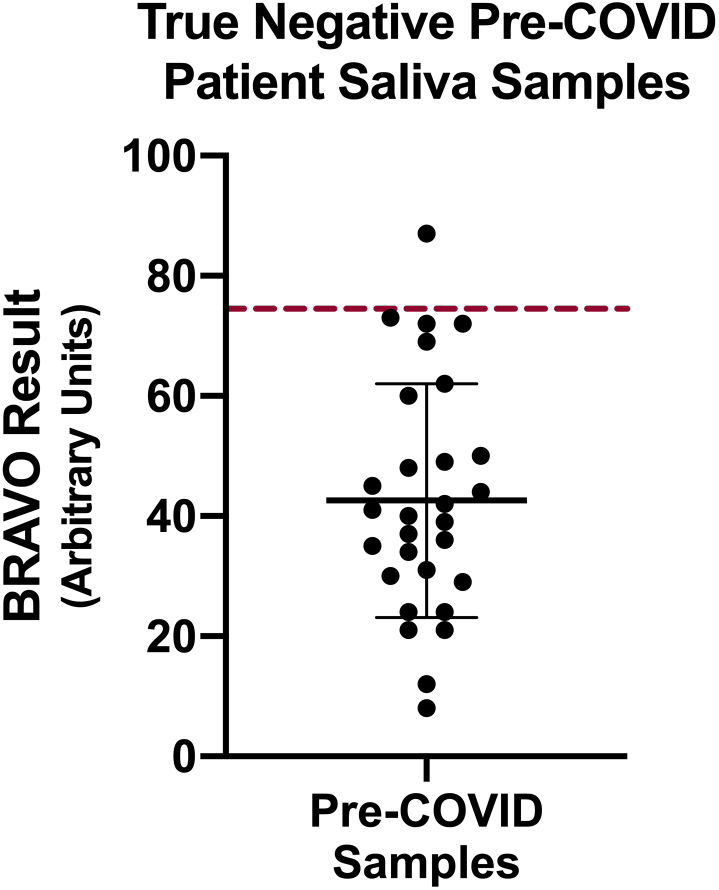
True negative (pre-COVID, acquired prior to December 2019) patient saliva samples were sourced from a commercial vendor and assessed using the BRAVO assay. Although variation in detectable salivary IgA existed across the samples (potentially due to polyreactive IgA or cross-reactivity with prior coronavirus exposure), the results clustered around a mean of 43.2 AU. We set a threshold cut-off to separate negative from positive samples where 95% of samples were negative. This value was set at 75 AU, or 1.65 standard deviations above the negative value mean (indicated by the broken red line).

We next investigated the potential for false-positive results from cross-reactivity with antibodies against other pathogens. We testing our assay against anti-sera against four common pathogens: anti-Influenza B IgG, anti-Respiratory Syncytial Virus IgG, anti-HIV, and anti-Hemophilus Influenza, as well as against Antinuclear Antibodies (ANA), nonspecific human IgG, and rheumatoid factors. Although we would have preferred to utilize patient-derived saliva samples, we were unable to acquire cross-reactive saliva samples, therefore we purchased commercially available patient-derived serum samples. Antibody concentrations in serum are 10-30x higher than concentrations typically found in saliva (IgA in saliva is 6.5-14.5 mg/dl^26^ while IgA in serum is 70-400 mg/dl^27^), therefore, before testing, we conservatively diluted the serum samples 10-fold using true-negative patient saliva sample as the diluent. We tested multiple independent samples from each potentially cross-reactive anti-serum, all of which were negative (**Table 1)**.

**Table 1.** BRAVO IgA Cross-Reactivity

| **Samples tested** | **Number**<br>**Samples** | **Positive (%)** |
| --- | --- | --- |
| Anti-Influenza B IgG | 5 | 0 (0%) |
| Anti-Respiratory syncytial virus IgG | 5 | 0 (0%) |
| Anti-HIV | 5 | 0 (0%) |
| Anti-Hemophilus influenza | 5 | 0 (0%) |
| Antinuclear Antibodies ANA | 3 | 0 (0%) |
| IgG | 5 | 0 (0%) |
| Rheumatoid Factors | 5 | 0 (0%) |

After validating and confirming our positive/negative threshold and demonstrating the lack of crossreactivity against common antibodies, we initiated clinical validation of samples from COVID-19 patients. We tested saliva from 38 patients who had previously tested PCR positive for SARS-CoV-2 PCR (**Figure 3**). 35 of the PCR-positive patients tested positive for salivary IgA against SARS-CoV-2, with a concentration reading above our threshold, while three tested negative. We therefore calculated a Positive Predictive Agreement (PPA; sensitivity) and a Negative Predictive Agreement (NPA; specificity) for our assay to be 92% and 97%, respectively (**Table 2**), with a p-value <0.0001 by two-tailed Fisher’s exact test.

**Figure 3.**
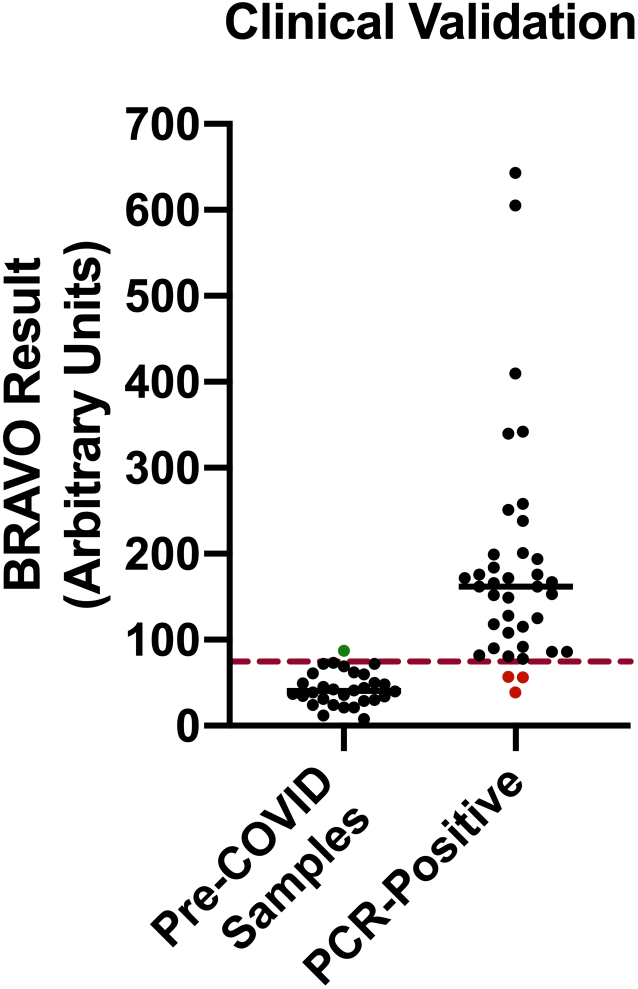
(A) The BRAVO clinical validation study included pre-COVID samples (collected prior to December 2019) as negative controls and saliva from PCR-confirmed COVID-19 patients as the positive control. The 30 negative control samples used to determine a cut-off for muco-positivity as described in the methods. One false positive (green dot) had readings above the cut-off. Of the 38 COVID-19 subjects, three (red dots) has salivary IgA levels below the cut-off and were deemed to be False Negative results.

**Table 2.** Results of Clinical Validation Study

|  | PCR Results/Clinical Truth |  |  |
| --- | --- | --- | --- |
| BRAVO Result |  | Positive | Negative |
|  | Positive | 35 | 1 |
|  | Negative | 3 | 29 |

Positive Percent Agreement = 92% (95% CI: 79.2, 97.3)
Negative Percent Agreement = 97% (95% CI: 83.3, 99.4)
p-value < 0.0001 by Fisher's Exact Test

To further characterize salivary muco-positivity, we tested saliva samples from an additional 53 high-risk subjects who had previously presented to Next Level Urgent Care Clinic (Houston, TX) with symptoms consistent with COVID-19. Information on their relevant history was collected from the subjects including information on COVID-19-related symptoms and any additional serology testing they may have undergone. 48 of the 53 additional subjects (91%) tested muco-positive for salivary IgA. The data from these 83 muco-positive subjects (48+35) was analyzed further.

As with the pre-COVID samples, we observed a broad range of salivary IgA concentrations (**Table 3**), with an over 9-fold difference between the extremes (78 and 720 AU). There was clear separation between the muco-positive and the pre-COVID samples, with means of 201 and 43 AU, respectively. Interestingly, the distribution of the muco-positive subjects shows what could be a couple of discontinuities in IgA levels. These could be a statistical artifact that will disappear with additional testing, or could have biological significance.

**Table 3.** Salivary IgA Levels in Muco-Positive Subjects (N=83)

|  | **Salivary IgA (AU)** |
| --- | --- |
| Mean | 201 |
| Median | 172 |
| Standard Deviation | 123 |
| Minimum | 78 |
| Maximum | 720 |

We then looked for correlations between IgA levels and other subject attributes. We examined the correlation between IgA levels and elapsed time between onset of symptoms and saliva collection (**Figure 4A**). For the ten subjects for whom we did not have information on the advent of symptoms, we used the PCR date. We lacked information on either date for one subject, who was excluded from the analysis. The elapsed time ranged from 32-109 days with a median of 61 days, with elevated salivary IgA observed over three months after symptom onset. We did not measure salivary IgA levels longitudinally in individual subjects, so we looked at other indicators of persistence. There was no correlation between elapsed time and IgA levels (r=0.015). Further, the average BRAVO result for samples collected before and after the median elapsed time was 194 and 207 AU, respectively (medians 167 and 176, respectively). These data suggest that mucosal IgA can persist for months, consistent with that reported for other respiratory diseases^28^.

**Figure 4.**
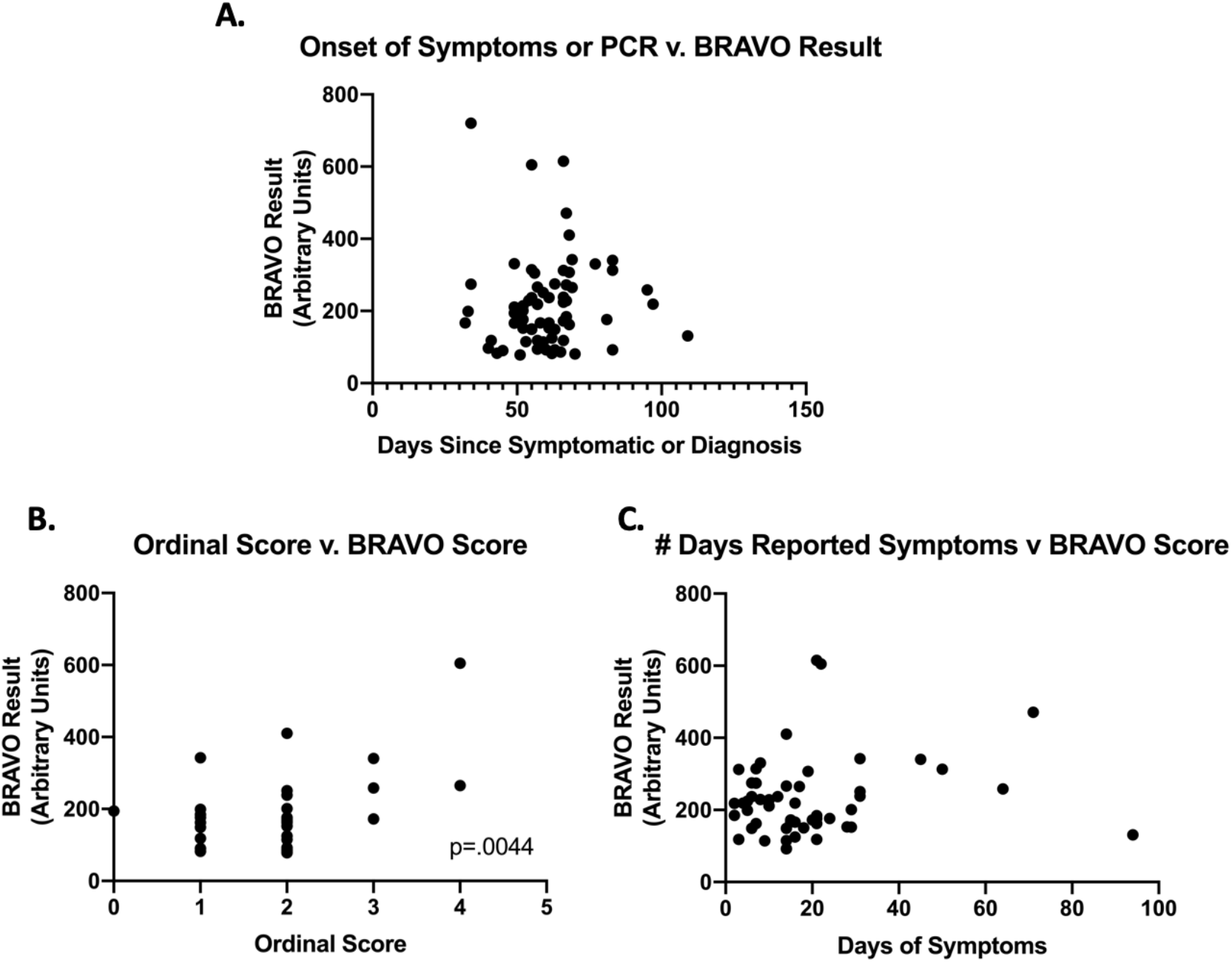
A) The number of days since onset of symptoms or the number of days since a positive PCR result (whichever was greater) was plotted against the BRAVO result. No correlation was identified. B) At the time of sample collection, patients were asked to rate their COVID-19 disease severity by using the WHO Ordinal Scale, where 0-4 indicates mild disease (increasing in severity) and >5 indicates severe disease. We observed a correlation between disease severity and BRAVO result, with a two-tailed Pearson Correlation of 0.4405 and a p-value=.0044. (C) For patients who were able to record the number of days of symptoms, we assessed if IgA levels (as determined via BRAVO score) correlated with duration. We did not identify any correlation.

As part of our clinical validation study, we further collected data regarding disease severity, according to the WHO Ordinal Scale for Clinical improvement, and symptom duration. There appeared to be a correlation with disease severity (r=0.446; p<0.01; **Figure 4B**). This is consistent with other reports on correlation between disease severity and serum antibody levels^29^. There was no apparent correlation between salivary IgA levels and duration of symptoms (r=0.17; **Figure 4C**)

To minimize risk to lab personnel of exposure to SARS-CoV-2, our clinical study was limited to salivary samples collected from individuals who were at least a month post-symptom onset, so we cannot report on when IgA levels first appear in saliva, though that work is currently underway. However, in limited anecdotal data collected since, we observe IgA levels arising early in the disease process, at the same time as with viral shedding. This early appearance of mucosal IgA is consistent with its role in protection against respiratory pathogens and is also consistent with reports that SARS-CoV-2 serum IgA appears earlier than IgG or IgM, possibly as early as two days after onset of symptoms.^22,23^ Interestingly, we also observed muco-conversion in several individuals despite a lack of symptoms, any evidence of disease (by PCR and/or serology) or known exposure to COVID-19. This is again anecdotal, but we are sharing it since it raises some intriguing questions.

Finally, we sought to collect additional data regarding the utility of saliva collection for this, and future assays. Patients and sample donors in our optimization and clinical validation studies were asked to complete a questionnaire about sample collection, including an estimation of the overall time required and an assessment of the ease of use for the self-collection. Saliva collection in this study was by supervised self-collection. Time required to collect the sample, measured by the supervisor, was 5-10 minutes in each case (**Table 4**). On a scale from 1-10 (with 1 indicating a high level of ease, and 10 indicating a high degree of difficulty), ~93% of donors selected 1 (indicating that the self-collection was very easy), ~5% selected 2, and roughly ~2% selected 3. No donors indicated any higher values (**Table 5**).

**Table 4.** Saliva sample collection time survey.

| **Survey choice** | **Number of respondents** |
| --- | --- |
| <5 minutes | None |
| 5-10 minutes | 87 |
| > 10 minutes | None |

**Table 5.** Salivary sample collection ease-of-use survey.

| **Score** | **Number of respondents (percent)** |
| --- | --- |
| 1 | 81 (93%) |
| 2 | 4 (5%) |
| 3 | 2 (2%) |
| 4-10 | 0 (0%) |
| Total | 87 |

## DISCUSSION

The classic model of human immunity comprises two interconnected, complementary systems: the innate immune system and the adaptive immune system. Lung mucosal immunity can act as a critical link between those systems with respiratory pathogens, playing an important, front-line defensive role. Lung mucosal immunity includes inducible innate resistance mediated by reactive oxygen species and antimicrobial peptides,^30^ as well as effectors - primarily secretory immunoglobulins - that are generated by and integrated with adaptive immunity. Mucosal immunity has been shown to play a crucial role against a broad range of respiratory pathogens, although its role in COVID-19 has not yet been established.

Secretory IgA (sIgA) is a principal component of mucosal immunity, and can easily be measured in saliva. Brevitest has validated a simple assay for measuring salivary IgA against SARS-CoV-2, which has been submitted to FDA for an Emergency Use Authorization (EUA). To our knowledge, no salivary IgA tests for SARS-CoV-2 have been authorized to date, and authorization of the BRAVO test will provide a new tool to assess immune responses in COVID-19. The Brevitest platform, on which our BRAVO assay runs, is designed to be a rapid (<15-minute), point-of-care, quantitative immunoassay. Although we have not yet validated the test for point-of-care (PoC) use, that work is currently underway. Given the convenience of saliva collection, a PoC test could open the door to expanded testing in multiple settings.

Here we demonstrate that quantitative data from true negative (pre-COVID) samples could be used to cleanly define a binary, qualitative threshold to assess muco-positivity. While the test we have submitted to FDA for an EUA is qualitative, the quantitative results we report here raise important questions about the host response to the SARS-CoV-2 virus, with potentially significant implications for the COVID-19 pandemic. These findings suggest that a more robust understanding of the role of mucosal immunity in COVID-19 could improve decision-making by patients, clinicians, and policymakers. We discuss some of these implications in three settings: individual immunity and clinical implications; community surveillance and herd immunity; and vaccine development.

### Individual Immunity and Clinical Implications

Our observations that (i) we see a large variation in salivary IgA titer, even in pre-COVID-19 samples; (ii) elevated IgA levels appear to persist for at least 2-3 months; and (iii) individuals may develop mucosal IgA without an overt SARS-CoV-2 infection, each raise intriguing questions.

Our unexpected detection of SARS-CoV-2 IgA antibodies in pre-COVID-19 saliva (albeit in significantly lower quantities than in confirmed COVID exposed patients) is supported by the literature. Polyreactive “natural” sIgA was first discovered in human saliva and colostrum^31^ and can bind to a number of pathogenic antigens. Polyreactive IgA appears to be a carryover from a primordial immune system that co-exists in humans along with the more modern adaptive immune system, and acts as a front-line host response that prevents epithelial entry of large numbers of microorganisms.^31,32^ Polyreactive IgA may be artifacts of prior adaptive responses, since IgA production - like IgG - generates memory cells that are stored for future use and expansion. These “natural” antibodies may display variable affinity against novel pathogens, as those pathogens may have epitopes in common with prior pathogens. However, these antibodies can also bind with affinities comparable to those seen with monoreactive antibodies. Further, other group have recently reported the presence of both cellular^33^ and humoral^34,35^ immunity to SARS-CoV-2 in pre-COVID samples, which have been attributed to prior exposure to other coronaviruses. We do not know if the salivary IgA antibodies detected in the pre-COVID-19 samples are protective, or if protection is related to mucosal antibody concentrations. But if these natural antibodies are protective, or accelerate a full adaptive response by giving the immune system an effective “seed” for subsequentrounds of somatic hypermutation and clonal selection, they could contribute to the variable susceptibility to COVID-19 infection that has been observed.

Intriguingly, even among the 83 muco-positive subjects in our study, we observed an over 9-fold range in IgA levels. If mucosal IgA in COVID-19 behaves in ways similar to those described for other respiratory pathogens, salivary IgA levels at either end of the spectrum (very low or very high) may have a clinical significance.

On the one hand, low levels of mucosal IgA have been associated with an increased incidence of respiratory infections. Byars et al. reviewed data from 1,189,061 individuals who had undergone tonsillectomy or adenoidectomy in childhood with a comparably sized control group, and found a significant increase in respiratory infections.^36^ Several groups have looked at correlation between changes in salivary IgA and respiratory infections in individuals and found that decreases in IgA correlated with an increase in infections. These findings have been reported in a wide variety of subjects including from elite athletes,^37^ infants,^38^ healthy children^39,40,41^ and children with Down’s syndrome.^42^ These findings suggest that individuals that are muco-negative for salivary IgA against SARS-CoV-2 may be at an elevated risk for infection and/or disease severity.

Conversely, there is also data suggesting that excessive levels of mucosal IgA can induce a pathologic inflammatory response. For example, both SIDS^43,44^ and “near-miss” SIDS^45^ infants had hyperimmune responses with substantially elevated levels of mucosal antibody responses. Yu et al. also reported a correlation between IgA levels and APACHE-II score in critically ill patients with COVID-19 (r^2^=0.72, P=0.01)^23^ although they did not examine salivary IgA. COVID-19 is characterized by sudden respiratory deterioration in patients, and sIgA can induce an inflammatory response in the airway, including neutrophil activation,^46^ and recent pre-prints suggest that systemic neutrophilic activation appears to be a marker of severity in pandemic flu^47^ and COVID-19.^48,49^ Thus, it may be desirable to look for correlations between clinical outcome and excess salivary IgA levels. If such a correlation is established, increased salivary IgA could serve as a biomarker to identify patients at an elevated risk of clinical deterioration in COVID-19, and perhaps candidates for early intervention with steroids, which have been shown to be effective in some ventilated patients.^50^

One of our intriguing, though anecdotal observations was that individuals may develop mucosal IgA without an overt COVID-19 infection. We have conducted over 250 assays, including those with earlier versions of the test. Brevitest employees, including several of the authors on this paper, contributed multiple saliva samples over the course of two months as the assay was refined. Several individuals muco-converted over this period of time despite no known exposure to COVID-19, no symptoms, and no evidence of disease by PCR and/or serology.

An important role played by sIgA is to continually scan the mucosal surface and prevent pathogen entry through “immune exclusion”^20^ where the polymeric sIgA binds to the pathogen and prevents it from binding to its cell surface targets until it is eliminated by clearance or degradation. If successful, immune exclusion prevents the pathogen from penetrating the epithelial defenses and establishing an infection, while still potentially triggering a ramp-up of the body’s immune defenses through their interaction with the adaptive immune system. Part of IgA’s function is to engage with a broad range of host cells. The FcaR receptor to the IgA Fc region is found on multiple cells including both key immune mediators like dendritic cells, and epithelial cells resulting in a broad range of effector functions involving both humoral and cellular adaptive responses. Once triggered, the adaptive humoral response produces both plasma cells and memory cells, and an active class-switching process.^28,32,51^ We need deeper understanding of the role played by mucosal IgA and its interplay with other elements of the immune system in order to better understand the pathogenesis and epidemiology of COVID-19.

### Community Surveillance and Herd Immunity

Reports that systemic IgA may be detectable earlier than IgG or IgM,^22,23^ as early as two days after symptom onset are consistent with the early-response role played by IgA, as well as with our anecdotal observations that individuals can muco-convert to positive salivary IgA contemporaneously with viral detection by PCR. A major challenge to successful containment of COVID-19 has been the relatively high false negativity in PCR testing as well as lack of timely results that have impeded contact tracing. If the early emergence of SARS-CoV-2 IgA in saliva is validated, salivary IgA may serve as a useful POC early screening tool, far more attractive to patients and clinicians than a nasopharyngeal swab, especially with children, with viral PCR serving as a confirmatory test.

While we do not know whether sIgA antibodies protect against SARS-CoV-2 exposure, extensive experience with other respiratory pathogens suggests that they should provide at least some protection. Our observation that mucosal IgA antibodies against SARS-CoV-2 persist for months is especially intriguing considering the recently reported data that systemic IgG antibody titers can decay within a month.^52^ We did not track individual IgA responses over time (that work is currently underway), but experience with a number of other respiratory pathogens suggests that mucosal IgA defenses can persist for extended periods of time.^15^

Apart from playing a special role in respiratory infections, salivary IgA also interacts with the adaptive immune system. Hence it is not surprising that, as with other respiratory infections, salivary IgA appears to correlate with systemic antibody levels against SARS-CoV-2.^24,25^ Given the simplicity of saliva collection, salivary IgA could be a particularly attractive tool for community surveillance including identifying and tracking vulnerable populations at increased risk. Sullivan et al. are currently validating home collection methods, including for salivary IgA.^53^

Moreover, if sIgA reduces the percentage of SARS-CoV-2 challenges that become full-blown COVID-19 infections, mucosal immunity could be part of the reason why there appears to be an unusually large number of asymptomatic patients who have tested positive for the virus. The virus could be present and neutralized by sIgA in mucosal tissue, and thus detectable by RT-PCR, but unable to enter epithelial cells in sufficient quantity to trigger the classic systemic immune response precipitated by a viral infection. And finally, if our data on muco-positivity rates are at least somewhat representative of the true community rate, and mucosal IgA is protective, we may be closer to herd immunity than currently believed.

### Vaccines and Mucosal IgA

Understanding whether mucosal IgA plays a protective role can help determine the trade-offs between topical and parenteral vaccines (or whether we might need both). Live attenuated flu vaccines have demonstrated encouraging activity,^54,55,56,57^ and several inhalation and intranasal vaccines for COVID-19 are under development. Especially intriguing is our anecdotal finding that exposure to sub-infective doses of SARS-CoV-2 could induce a mucosal immune response as measured by salivary IgA. A speculative interpretation is that the threshold of viral exposure required to induce a mucosal IgA response may be below that required to induce and establish infection or generate a measurable IgG response. Could we already have an effective and scalable live attenuated vaccine, with attenuation done by dose-reduction to a non-infective titer?

Even with parenteral vaccines, tracking salivary IgA may be useful for several reasons. Not all mucosal vaccines lead to systemic immunity, but many do.^57,58,59,60,61,62^ Selecting a vaccine that induces both mucosal and systemic immunity in a Phase 1 clinical study might increase the probability of success in Phase 3. Conversely, high levels of salivary IgA have been associated with a blunted response to immunization,^54^ and over-representation of subjects with elevated sIgA in a placebo arm may mask the beneficial effect of a vaccine. Thus, it may be prudent to analyze or even stratify subjects in a vaccine clinical study by salivary IgA levels.

In summary, the preliminary data on salivary IgA serology that we have generated with the Brevitest BRAVO assay is consistent with the extensive published data on the critical role played by mucosal immunity in protecting against respiratory viruses. This makes a strong case for the clinical, research, and public health communities to better understand sIgA’s role in COVID-19 and to evaluate the possibility of its use in patient management, community monitoring, and vaccine development.

## METHODS

### Brevitest Platform and BRAVO Assay

The Brevitest platform consists of an Analyzer and a single-use test Cartridge. The Analyzer is a miniaturized fully automated robotic ELISA device that can conduct a standard ELISA using 40 microliters of sample in less than fifteen minutes. BRAVO is an enzyme-Linked Immunosorbent Assay (ELISA) capable of quantitative detection of IgA antibodies to SARS-CoV-2 in human saliva though the assay has only been validated for qualitative IgA detection. The analyzer controls and automates the ELISA process. The automated mechanism controls the movement of magnetic microspheres across several compartments within the cartridge. In each compartment a separate reaction takes place on the surface of these microspheres. The microspheres are oscillated in each compartment using proprietary mixing patterns. The Analyzer incorporates a scanner that measures the optical signal at the end of the reaction and sends the data to the cloud for analysis. There are a number of controls and safeguards built into the platform to ensure that the cartridge is inserted properly, the correct test is performed, the cartridge is not past its expiry date, that the results are valid, and that patient identifiable information is not at risk.

The cartridges are loaded with about 40μl of patient’s diluted saliva (diluted 5x in phosphate-buffered saline), and inserted into the analyzer. The analyzer is a cloud connected robotic device which moves the magnetic microspheres within the cartridge through the different compartments to perform the complete ELISA reaction. The color change at the end of the process is read by the analyzer’s on-board optical sensors. BRAVO uses the following reagents. Capture antigen: 1:1 ratio of SARS-CoV-2 Nucleocapsid antigen and SARS-CoV-2 Spike S1 antigen (Receptor Binding Domain), both purchased from Genscript. Secondary antibody: Goat Anti-Human IgA conjugated to HRP (Cat: PA1-74395, Thermo Fisher Scientific). COVID-19 antibody (Absolute Antibody) as our internal positive control.

Each BRAVO assay utilizes a single-use cartridge, which in addition to the sample lane, contains separate positive- and negative-control lanes to ensure consistency. The optical density in the control lanes serve as a quality check on the assay/cartridge and are also used to inform the sample reading. The BRAVO result is determined by an algorithm that incorporates the results from the sample lane as well as the negative and positive controls, thereby controlling for cartridge to cartridge variation as well as any minor differences in the ambient environment.

### Assay Validation

The BRAVO assay was validated as described in the Results section, using true negative pre-COVID saliva samples (Lee Biosolutions) and tested for cross-reactivity using serum samples for anti-influenza B IgG, anti-respiratory syncytial virus IgG, antinuclear antibodies, IgG, rheumatoid factors (Biomex ZMBH), HIV, and Hemophilus influenza antibodies (Cantor Bioconnect).

### Participant Enrollment and Sample Collection

For clinical sample collection, we enrolled participants who had presented themselves with symptoms consistent with COVID-19 infection to the Next Level Urgent Care Clinic (Houston, TX) and subsequently underwent a PCR test using nasopharyngeal sample collection. Information on relevant history was collected from the subjects including information on COVID-19-related symptoms and any additional testing (PCR or serology) they may have undergone. Subjects were also asked to rate the ease of saliva sample collection on a scale of 1 (easy) to 10 (difficult). The details of the PCR test used for each subject were obtained from the clinic’s electronic health records. The clinical protocol for sample and data collection and the informed consent document were approved by IntegReview, an independent Institutional Review Board.

For saliva collection, subjects were provided with a kit containing a salivary swab, a plastic syringe (Care Touch, part number CTSLL5) and a 1.5 ml capacity tube (VWR International, part number 20170-577). The swabs were FDA listed Class 1 saliva collection devices, safe for use in adults or children 6 years or older, and are qualified for recovery of analytes, comparable to saliva samples collected from passive drool. All three components were supplied individually wrapped by the manufacturer, and assembled in a single bag by Brevitest. Each bag contained a syringe, a collection tube and a salivary swab, together with instructions for saliva collection.

For our clinical validation, subjects self-collected saliva supervised by a trained provider. The individuals supervising the procedure completed a survey as to the length of time taken for sample collection (<5 minutes, 5-10 minutes, or >10 minutes). Individuals also responded rating the procedure on a scale of 1 (easiest) to 10 (most difficult).

The collection procedure is briefly described here. Specimens were collected in accordance with correct medical practices, observing all standard safety procedures for both patient and collector. The subjects were briefly verbally instructed on the procedure and a glove put on their dominant hand. They then opened the bag containing the swab package, syringe and collection tube, removed the contents, and released the swab from its packaging. The swab was placed under the tongue and kept there for 2 minutes. Meanwhile, the subject identifier code label was put on the tube, the syringe removed from its package and the plunger removed from the barrel. After collection, the swab was placed in the barrel, followed by the plunger. The tip of the syringe was inserted into the collection tube and the plunger slowly pushed down to squeeze the collected saliva into the tube. The tube was then closed and the user confirmed that the volume of saliva collected (excluding the bubbles/froth on top) reached the first mark on the tube (which ensures 250 μl of sample). If the sample volume was inadequate, the procedure was repeated, with the second sample added to the tube to make up the required volume. The closed tube was placed in a biosafety pouch by the healthcare provider and put in a freezer or container with dry ice for storage below -20°C and later transport. Samples were kept frozen until actual testing, for a minimum of 2 hours at -20°C.

### Saliva sample preparation

The frozen samples in collection tubes were thawed and warmed up to room temperature by removal from the freezer 10 to 15 minutes prior to testing. 400 μl of PBS (phosphate-buffered saline) was placed in a new separate 1.5 ml tube. 100 μl of the saliva sample was added to this tube, and mixed with the PBS by gently inverting the tube 3-5 times. Care was taken to take the saliva sample from the bottom of the sample tube, avoiding any remaining bubbles or froth at the top.

### Cartridge loading and code linking

An appropriate cartridge for the BRAVO assay was taken from the refrigerator, removed from its pouch and placed on a clean, flat, dry surface with the barcode on the cartridge facing up. Before running the assay, the sample was electronically connected to the cartridge, to enable automated reading of results after the test. To electronically link the sample identification barcode (on the collection tube) to the cartridge code (on the cartridge), any web-enabled device (e.g. tablet or smartphone) with a camera and web browser could be used. The test operator logged into the appropriate website on the device and sequentially scanned the code on the cartridge, followed by the subject identifier code on the tube. Finally the linking (if done satisfactorily) was saved and the cartridge was ready to be used. 40 μl of the diluted sample was pipetted into the sample well in the cartridge using a calibrated pipette. The rest of the sample was saved until the test run was complete, in case of failures or the need to re-test.

### Statistical analysis

The PPA and NPA were calculated using standard procedures as explained in the Results section. The confidence limits were calculated for the population proportion using the Wilson method with standard statistical software. Inputs were the sample size and number of positive results, the desired level of confidence in the estimate (95%) and the number of decimal places required in the answer. Outputs were the estimated proportion plus upper and lower limits of the specified confidence interval.

## Data Availability

Individual subject-level data included in the manuscript.

## Competing Interests

AV, DC, JG, PG, AL, CF, LL are employees of BreviTest Technologies LLC., and Fannin Innovation Studio.

JD is an intern at BreviTest Technologies, LLC.

## Research Support

Development of the Brevitest platform was supported by the National Institute on Drug Abuse (NIDA) and the National Institute on Minority Health and Health Disparities (NIMHD) at the National Institutes of Health (NIH), Grant Numbers: 1 R43 DA041966-01; 1 R43 MD013409-01A1; 1 R43 DA043325-01, 2 R44 DA043325-02, 5 R44 DA043325-03; 3 R43 DA041966-01S1, the Vijay and Marie Goradia Foundation, and Fannin Innovation Studio.

## Author Contributions

All authors made contributions to the conception, design, or execution of the work. All authors have reviewed the manuscript critically and have given final approval to the manuscript.

## Notes

### Clinical Trial

Clinical trial protocol was used for sample collection. No subjects were treated.

### Author Declarations

The protocol and informed consent form for collection of saliva samples for this Minimal Risk Study received expedited approval by IntegReview IRB, Austin, TX.

